# Clinical Trial of Efficacy and Toxicity of Disoproxil Tenofovir Fumarate and Emtricitabine for Mild to Moderate SARS-CoV-2 Infections

**DOI:** 10.1101/2021.09.28.21264242

**Authors:** E.A.G. Arruda, R.J. Pires-Neto, M.S. Medeiros, J. Quirino-Filho, M. Clementino, R.N.D.G. Gondim, L.M.V.C. Magalhães, K.F. Cavalcante, V.A.F. Viana, Liana Perdigão Mello, R.B Martins, A.A. Santos, P.J.C. Magalhães, A. Havt, N.P. Lopes, E. Arruda-Neto, A.A.M. Lima, study group members

## Abstract

This study aimed to evaluate the efficacy and toxicity of tenofovir (TDF) and TDF combined with emtricitabine (TDF/FTC) in patients with mild to moderate COVID-19 infections. We conducted a randomized, double-blind, placebo-controlled clinical trial in patients with clinical suspicion of mild to moderate respiratory infection caused by SARS-CoV-2 who were treated at an outpatient clinic. Patients were randomly recruited to take 10 days of TDF (300 mg/day), TDF (300 mg/day) combined with FTC (200 mg/day) or placebo Vitamin C (500 mg/day). The primary parameter was the score of symptoms and predictive signs of COVID-19, assessed on the seventh day of patient follow-up. From a total of 309 patients with clinical suspicion of SARS-CoV-2, 227 met the inclusion criteria and were randomly distributed into the following groups: (a) 75 (one did not initiate treatment) in the TDF group; (b) 74 in the TDF combined with FTC group; and (c) 77 in the Vitamin C group (placebo). Of the 226 patients, 139 (62%) were positive for SARS-CoV-2. Fever (≥37.8°C), ageusia or dysgeusia, anosmia or dysosmia, and two or more clinical symptoms or signs were significantly associated with SARS-CoV-2 infection. There was no significant change in clinical score based on clinical symptoms and signs between treatment groups. Patients with mild to moderate infection by SARS-CoV-2 had higher concentrations of G-CSF, IL-1β, IL-6 and TNF-α compared to patients without infection. Patients with mild to moderate respiratory infection, with fever (≥37.8°C), loss of smell, loss of taste and two or more symptoms, have a better prediction for the diagnosis of COVID-19. Patients with SARS-CoV-2 showed higher and more persistent proinflammatory cytokines profile compared to patients not infected with SARS-CoV-2. Pharmacological intervention with TDF or TDF combined with FTC did not change the clinical signs and symptoms score in mild to moderate respiratory infection in patients with SARS-CoV-2 compared to the Vitamin C group (placebo).

## Introduction

The signs and symptoms of COVID-19 are sometimes difficult to differentiate from other viral respiratory infections. Defined clinical predictors for the new coronavirus (SARS-CoV-2) can help guide timely therapy and develop clinical score for research antiviral therapy, as well as avoid unnecessary antibiotics, guide proper isolation and prevention of COVID transmission-19^1-3^. The evaluation of inflammatory biomarkers such as cytokines, chemokines, and cell growth factors in the pathobiology of mild to moderate infection of COVID-19 can help in better knowledge of the disease, treatment decisions, prognosis and in the development of therapies for prevention and treatment. Several therapeutic agents have been evaluated for the treatment of COVID-19, but only the antiviral drug called remdesivir, administered intravenously, has shown efficacy in reducing the duration of the disease by 26.7% in critically ill patients^4^. No data on the efficacy and toxicity of tenofovir disoproxil fumarate (TDF) in coronovirus infections are available to date^5^. Recent studies on antiretroviral drugs with actions against the coronovirus have suggested promising results in the specific inhibition of SARS-CoV-2 polymerase (RNA-dependent RNA polymerase: RdRp) with drugs such as tenofovir (TDF) and emtricitabine (FTC) in addition to remdesivir^6-8^. In silico studies have shown that TDF has a coupler binding to the RdRp of SARS-CoV-2 and causes inhibition of infection *in vitro* in cell cultures^6,7^. Furthermore, the TDF/FTC combination reduced the viral load in nasopharyngeal samples from animals (ferrets) experimentally infected with SARS-CoV-2^8^. These results led us to evaluate TDF alone or TDF combined with FTC in reducing the clinical duration, observing the signs and symptoms of COVID-19 evaluated in the 7th. day of the experimental protocol. This study evaluated the efficacy and toxicity of TDF alone or in combination with FTC in a randomized, double-blind, placebo-controlled clinical trial on the duration of COVID-19 through clinical observation of the onset and end of disease signs and symptoms. In addition, we evaluated biomarkers of cytokines, chemokines, and cell growth factors associated with mild to moderate respiratory infection with SARS-CoV-2 when compared to patients without SARS-CoV-2 infection.

## Material and methods

### Study design, site, and population

The study design was a prospective, clinical trial, randomized, double-blind, placebo-controlled. The study protocol and consent form were approved by CONEP with registration no. 34 18262.00.0000.5045 and the ClinicalTrials.gov Identifier record: NCT04712357. The study was carried out in the city of Fortaleza-CE, Brazil, with 2,686,607 inhabitants and which registered, until August 13, 2021, a total of 254,898 cases and 9,616 deaths from COVID-19, with a mortality rate equal to 357.9 per 100 thousand inhabitants^9^. After signing the consent form, patients with mild to moderate respiratory infection and clinical suspicion of COVID-19 were invited to participate in the study.

### Selection and recruitment of participants

We selected adult patients aged 18-60 years and of both genders, who met the following inclusion criteria: (a) age between 18-60 years; and (b) clinical suspicion of mild to moderate COVID-19 respiratory infection. Mild to moderate respiratory infection was defined as absence of dyspnea, respiratory distress and O_2_ saturation < 95%. Exclusion criteria were: (a) the patient was already receiving some study drug; (b) plan for hospitalization within the next 24 hours; (c) contraindication for the use of study drugs; (d) diagnosis of HIV and/or hepatitis B virus infections; (e) pregnant woman; and (f) patient with fixed residence outside the study municipality. The recruitment of patients took place from November 9, 2020, to July 5, 2021. In these eight months, a total of 309 patients were eligible for the study, 82 of which were excluded based on the inclusion and exclusion criteria. Thus, 227 patients were randomly distributed into the three treatment groups, using a random list in blocks of three sequentially permuted numbers. However, one patient did not start treatment. Thus, 75 patients took TDF (one patient did not initiate treatment; 300 mg/day), 74 patients took TDF (300 mg) combined with FTC (200 mg) and 77 patients took Vitamin C (500 mg/day), all for 10 days.

### Study duration and intervention regimen

The patients were followed up by the medical team and other health professionals during four visits: day 1, day 7 (±3 days), day 14 (±3 days) and day 28 (±3 days), at the outpatient clinic of Hospital São José de Infectious Diseases of the Ceará State Health Department, including the initial recruitment visit. On the day of recruitment, demographic and clinical data, nasopharyngeal swab samples, blood were collected, and information on adverse events and serious adverse events was reported, in addition to the delivery of study intervention drugs and initiation of treatment. On day 7, second visit, we collected clinical information, nasopharyngeal swab and blood samples, follow-up adverse event and serious adverse event information. The same data were collected on subsequent visits: third visit (day 14) and fourth visit (day 28).

### Molecular diagnosis of SARS-CoV-2

Molecular diagnosis was performed by Real-Time PCR (RT-qPCR) based on US CDC guidelines^10^. Nasopharyngeal swabs followed for nucleic acid (NA) inactivation and isolation and RT-qPCR uniplex. was made with commercially obtained primers and probes for SARS-CoV-2 (IDT, Newark, NJ - USA), in reactions of 20 uL with 10 uL of RT-PCR enzyme; 0.4 µl RT (Promega, Madison, WI - USA); 1.5 uL of primer/probe mix; 3.1 uL of autoclaved ultrapure water; and 5 µl of the RNA sample.

### Biomarkers in the immune-inflammatory response

IgM and IgG tests were performed with the LIAISON® SARS-CoV-2 S1/S2 IgG and IgM kit (DiaSorin, Saluggia, Italy) a chemiluminescence-based immunoassay for the quantitative determination of antibodies SARS-CoV-2 anti-S1 and anti-S2 IgG and qualitative IgM antibodies to SARS-CoV-2 in human serum or plasma samples, following the manufacturer’s protocol. For the evaluation of biomarkers of the inflammatory response, we used XMAP Luminex technology (Merck, Kenilworth, New Jersey), including the simultaneous analysis in the same serum sample (volume of 25 uL), of a panel of protein markers of pro and anti-inflammatory and cell growth factors (IL-6, MCP-3, IL-1β, IL1-RA, IL-10, G-CSF, TNF-α, MCP-1, IFN-γ, IP-10), following protocol manufacturer, with results expressed in pg/mL.

### Adverse events and serious adverse events system

Patients involved in the study were monitored for any Adverse Event (AE) and Severe Adverse Event (SAE). The SAEs were registered in a specific form, and they were reported within 14 days to the Study Management Group to the Research Ethics Committee (COMEPE). At the time of the evaluation, if signs and symptoms of worsening of the disease were observed, the patient was referred for return to medical care at the Hospital São José emergency. Adverse event (AE) was defined as any unpleasant medical occurrence that arose during the administration of the drugs clinical trial, with or without a causal relationship to the study agent. A serious adverse event (SAE) was defined as any adverse experience that occurred during the use of any of the study drugs, resulting in the following outcomes: death; threat to life; need for hospital admission; persistent or significant disability or disability; or any major medical event. All AEs and SAEs recorded were reviewed and reported according to good clinical practice procedures^11^. AEs and SAEs occurring during the study were reported in accordance with the reports and guidelines required by the US Division of Microbiology and Infectious Diseases for EAGs^12^ as well as to the Federal University Review Board of Ceará.

### Sample size and statistical analysis

The estimated sample size was calculated at 73 patients for each intervention group (Total = 219 patients) to achieve statistical power of 80% (1 – Beta; type-II error) and statistical significance of p = 0.05 (Alpha: type-I error) to reduce the clinical duration of the disease in the groups receiving the drugs by 20% compared to the clinical duration of the disease in the Vitamin C placebo control group, with 15% losses by exit from the study. For the calculation of the sample size, we used evidence-based clinical decision from ClinCalc.com for medical professionals (https://clincalc.com/Stats/SampleSize.aspx). Data were entered into spreadsheets and verified by 2 independent researchers to ensure accuracy. All data were de-identified and statistical analysis was done using SPSS Statistics 20.0 (IBMCorporation, https://www.ibm.com). We used the Shapiro-Wilk Test to assess the quantitative normality of the data and the Levene Test to assess the equality of variations. For non-parametric variables, the Mann-Whitney U Test, Kruskal-Wallis Test for independent samples or Wilcoxon Signed Rank Test for paired samples was used. Qualitative variables were used the χ^2^ test or Fisher’s exact test. We used GraphPad Prism version 3.0 for Windows (GraphPad Software, https://www.graphpad.com) and ArcGIS software version 9.0 (ESRI Co. Redlands, CA) were used for supplementary statistical analysis, table formatting and figure creation. Unadjusted and adjusted multivariate logistic regression was used to assess clinical signs and symptoms associated with SARS-CoV-2 infection and determination of clinical scores. Relative risk with 95% confidence intervals (CIs) was used to assess signs and symptoms associated with risk of SARS-CoV-2 infection. All statistical tests were 2-sided with a significance level of p < 0.05.

## Results

**Figure 1** shows the geographic location of the city of Fortaleza on the map of Brazil and the heat map of the dispersion of confirmed cases (25,683 georeferenced cases) at the peak of the second wave of COVID-19 in the city, in March 2021. This period coincides with the period of greatest recruitment in the clinical trial, under the predominance of the gamma variant of SARS-CoV-2. There was a large continuous agglomeration that occupied several neighborhoods such as Benfica, Damas, Bom Futuro, José Bonifácio and Parquelândia, close to Hospital São José, location of the source of recruitment of patients in the study. The map shows the spread of cases to the south of the metropolitan region of Fortaleza, CE, Brazil. The dots marked in black on the map represent the origin of patients recruited in the clinical trial, from November 9, 2020, to July 5, 2021. Demographic and comorbidity data of patients in the study are summarized in **Table 1**. The study patients enrolled in the intervention with TDF and TDF/FTC with clinical suspicion of mild to moderate COVID-19 were adults with a mean age of 38 years (with a standard deviation of 14.9); predominantly female; with the majority reporting having completed high school and university level. They were seen in the first 4 days after the onset of clinical symptoms and signs of the disease and had a median of 6 clinical symptoms and signs. Thirty-three percent reported one or more comorbidities, such as hypertension (17%), type 2 diabetes (10%), smoking (7%) and asthma (6%), among others less frequently (≤4%). These characteristics were not different between study groups. RT-qPCR results on nasopharyngeal swab samples showed that 62% (139) of the total number of patients recruited (226) were positive for SARS-CoV-2. Viral load in groups designed for intervention did not show significant differences at entry into the study protocol.

**Figure 1.**
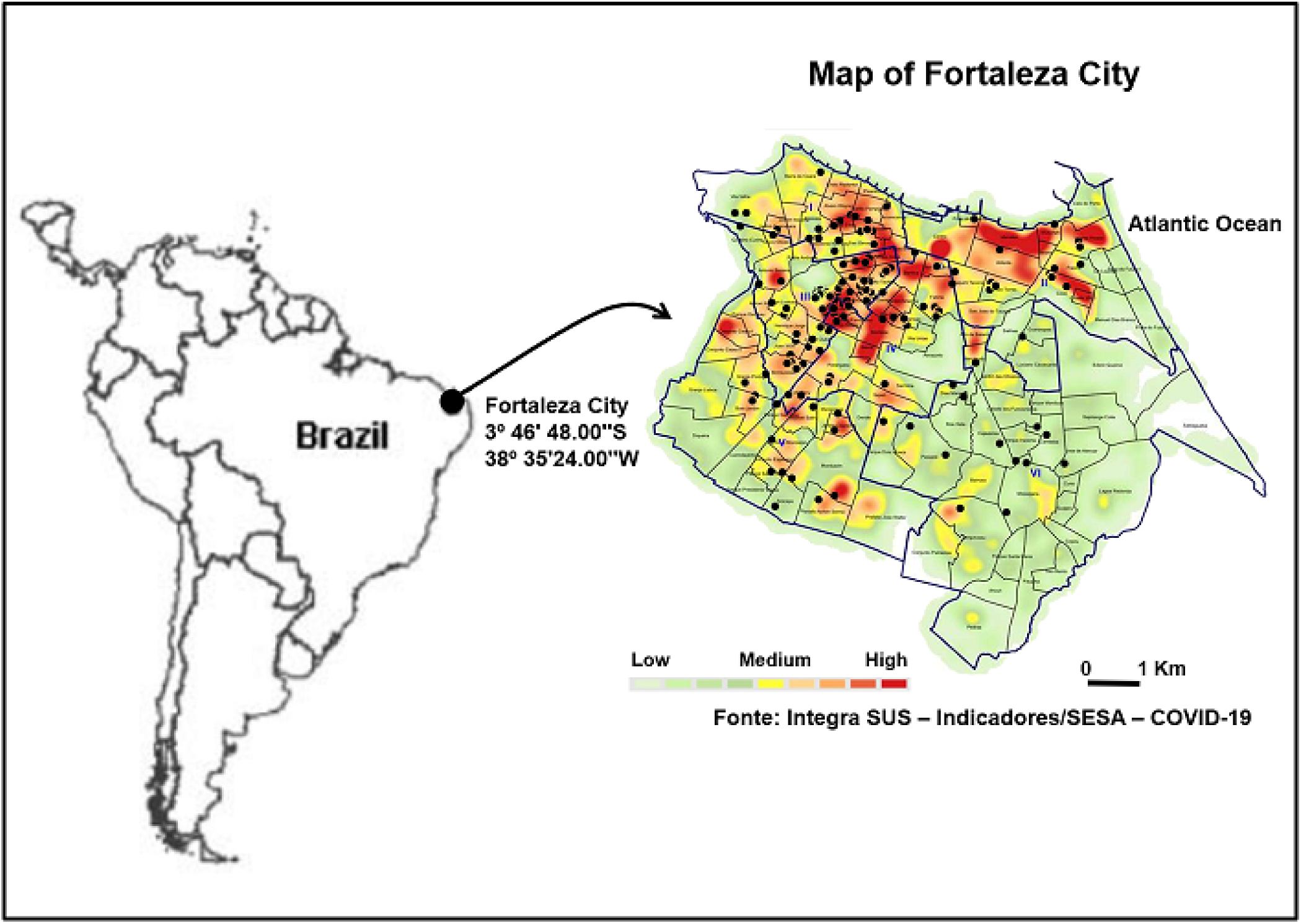
Map of Fortaleza city, state of Ceará, Brazil and study location. This figure shows the geographic location of the city of Fortaleza on the map of Brazil and the heat map of the dispersion of confirmed cases (25,683 georeferenced cases) at the peak of the second wave of COVID-19 in the city, in March 2021. This period coincides with the period of greatest recruitment in the clinical trial, under the predominance of the gamma variant of SARS-CoV-2. There was a large continuous agglomeration that occupied several neighborhoods such as Benfica, Damas, Bom Futuro, José Bonifácio and Parquelândia, close to Hospital São José, location of the source of recruitment of patients in the study. The map shows the spread of cases to the south of the metropolitan region of Fortaleza, CE, Brazil. The dots marked in black on the map represent the origin of patients recruited in the clinical trial, from November 9, 2020, to July 5, 2021.

**Table 1.**
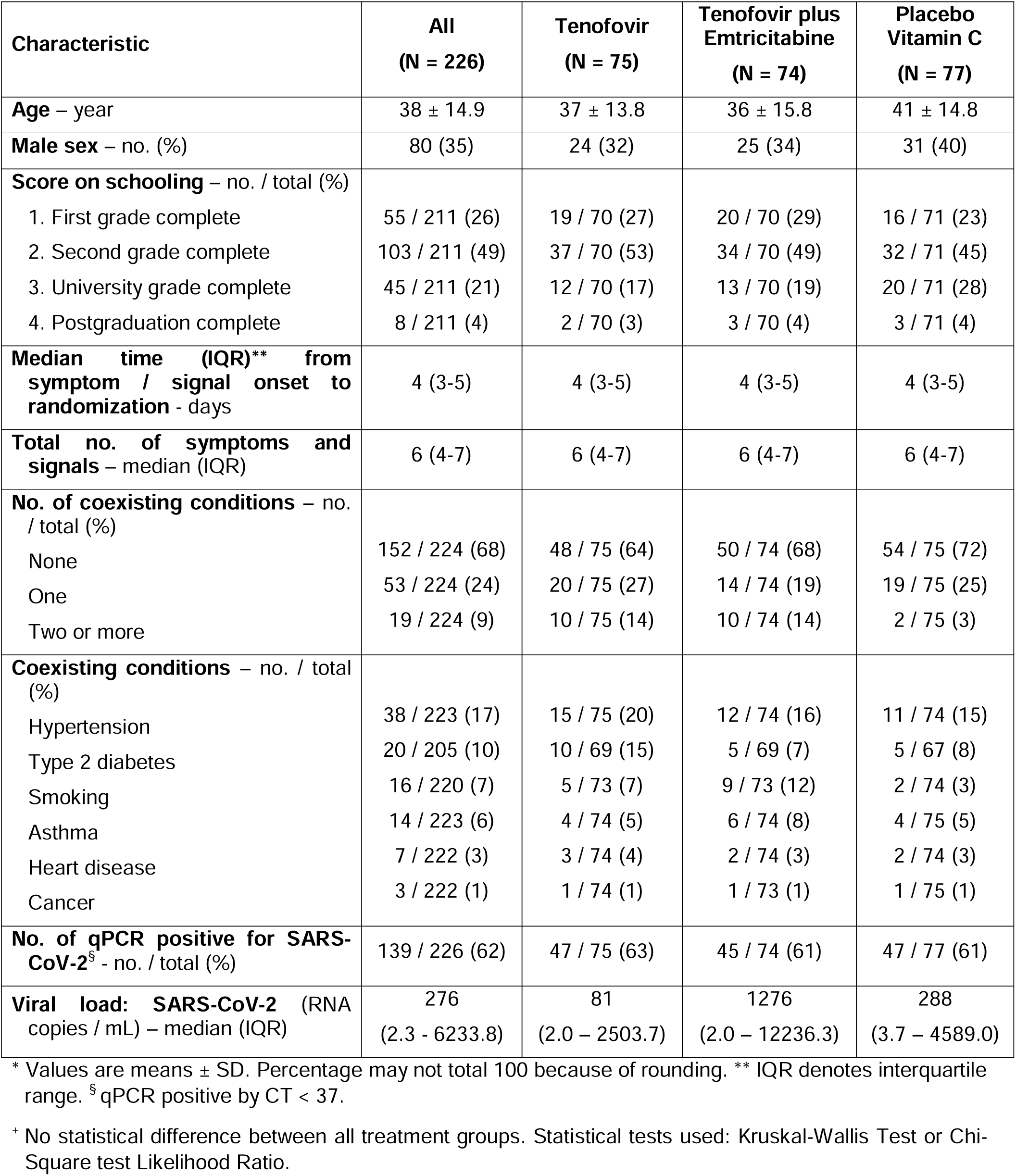
Demographic and clinical characteristics of the patients at baseline. ^*+^.

| **Characteristic** | **All**<br>**(N = 226)** | **Tenofovir**<br>**(N = 75)** | **Tenofovir plus**<br>**Emtricitabine**<br>**(N = 74)** | **Placebo**<br>**Vitamin C**<br>**(N = 77)** |
| --- | --- | --- | --- | --- |
| **Age – year** | 38 ± 14.9 | 37 ± 13.8 | 36 ± 15.8 | 41 ± 14.8 |
| **Male sex – no. (%)** | 80 (35) | 24 (32) | 25 (34) | 31 (40) |
| **Score on schooling – no. / total (%)** |  |  |  |  |
| 1. First grade complete | 55 / 211 (26) | 19 / 70 (27) | 20 / 70 (29) | 16 / 71 (23) |
| 2. Second grade complete | 103 / 211 (49) | 37 / 70 (53) | 34 / 70 (49) | 32 / 71 (45) |
| 3. University grade complete | 45 / 211 (21) | 12 / 70 (17) | 13 / 70 (19) | 20 / 71 (28) |
| 4. Postgraduation complete | 8 / 211 (4) | 2 / 70 (3) | 3 / 70 (4) | 3 / 71 (4) |
| **Median time (IQR)\*\* from**<br>**symptom / signal onset to**<br>**randomization - days** | 4 (3-5) | 4 (3-5) | 4 (3-5) | 4 (3-5) |
| **Total no. of symptoms and**<br>**signals – median (IQR)** | 6 (4-7) | 6 (4-7) | 6 (4-7) | 6 (4-7) |
| **No. of coexisting conditions – no.**<br>**/ total (%)** |  |  |  |  |
| None | 152 / 224 (68) | 48 / 75 (64) | 50 / 74 (68) | 54 / 75 (72) |
| One | 53 / 224 (24) | 20 / 75 (27) | 14 / 74 (19) | 19 / 75 (25) |
| Two or more | 19 / 224 (9) | 10 / 75 (14) | 10 / 74 (14) | 2 / 75 (3) |
| **Coexisting conditions – no. / total**<br>**(%)** |  |  |  |  |
| Hypertension | 38 / 223 (17) | 15 / 75 (20) | 12 / 74 (16) | 11 / 74 (15) |
| Type 2 diabetes | 20 / 205 (10) | 10 / 69 (15) | 5 / 69 (7) | 5 / 67 (8) |
| Smoking | 16 / 220 (7) | 5 / 73 (7) | 9 / 73 (12) | 2 / 74 (3) |
| Asthma | 14 / 223 (6) | 4 / 74 (5) | 6 / 74 (8) | 4 / 75 (5) |
| Heart disease | 7 / 222 (3) | 3 / 74 (4) | 2 / 74 (3) | 2 / 74 (3) |
| Cancer | 3 / 222 (1) | 1 / 74 (1) | 1 / 73 (1) | 1 / 75 (1) |
| **No. of qPCR positive for SARS-**<br>**CoV-2<sup>§</sup> - no. / total (%)** | 139 / 226 (62) | 47 / 75 (63) | 45 / 74 (61) | 47 / 77 (61) |
| **Viral load: SARS-CoV-2 (RNA**<br>**copies / mL) – median (IQR)** | 276<br>(2.3 - 6233.8) | 81<br>(2.0 – 2503.7) | 1276<br>(2.0 – 12236.3) | 288<br>(3.7 – 4589.0) |

\* Values are means ± SD. Percentage may not total 100 because of rounding. \*\* IQR denotes interquartile range. <sup>§</sup> qPCR positive by CT < 37.
<sup>†</sup> No statistical difference between all treatment groups. Statistical tests used: Kruskal-Wallis Test or Chi-Square test Likelihood Ratio.

The selection, recruitment and follow-up visits of patients are shown in **Figure 2**. Three hundred and nine patients with clinical suspicion of COVID-19 were selected and provided consent to record the demographic data and verify the inclusion and exclusion criteria for recruitment in the study. Eighty-two patients were excluded from the study for not meeting the inclusion criteria. Two hundred twenty-seven patients were randomized to receive one of three treatments: TDF (76 patients); TDF/FTC (74 patients) and Vitamin C placebo (77 patients). Only one patient in the TDF group did not start the medication, so 75 patients in this group started the treatment. At the first visit, 226 patients were treated for demographic and clinical data collection, nasopharyngeal and blood swab samples collected, implementation of the surveillance system for adverse events (AEs) and serious adverse events (SAEs), in addition to drug delivery and guidance to start the treatment. At the second visit (7 ± 3 days), 187 patients (83%) were seen to monitor clinical symptoms and signs, collect nasopharyngeal swab and blood samples, monitor AEs and SAEs, and check the medication taken during the period. At the third visit (14 ± 3 days), 155 patients (69%) attended to monitor clinical symptoms and signs, collect nasopharyngeal swab and blood samples, monitor AEs and EAGs, and check the medication taken during the period. On the fourth and last visit (28 ± 3 days), 129 patients (57%) were seen to monitor clinical symptoms and signs, collect nasopharyngeal swab and blood samples, and monitor AEs and SAEs.

**Figure 2.**
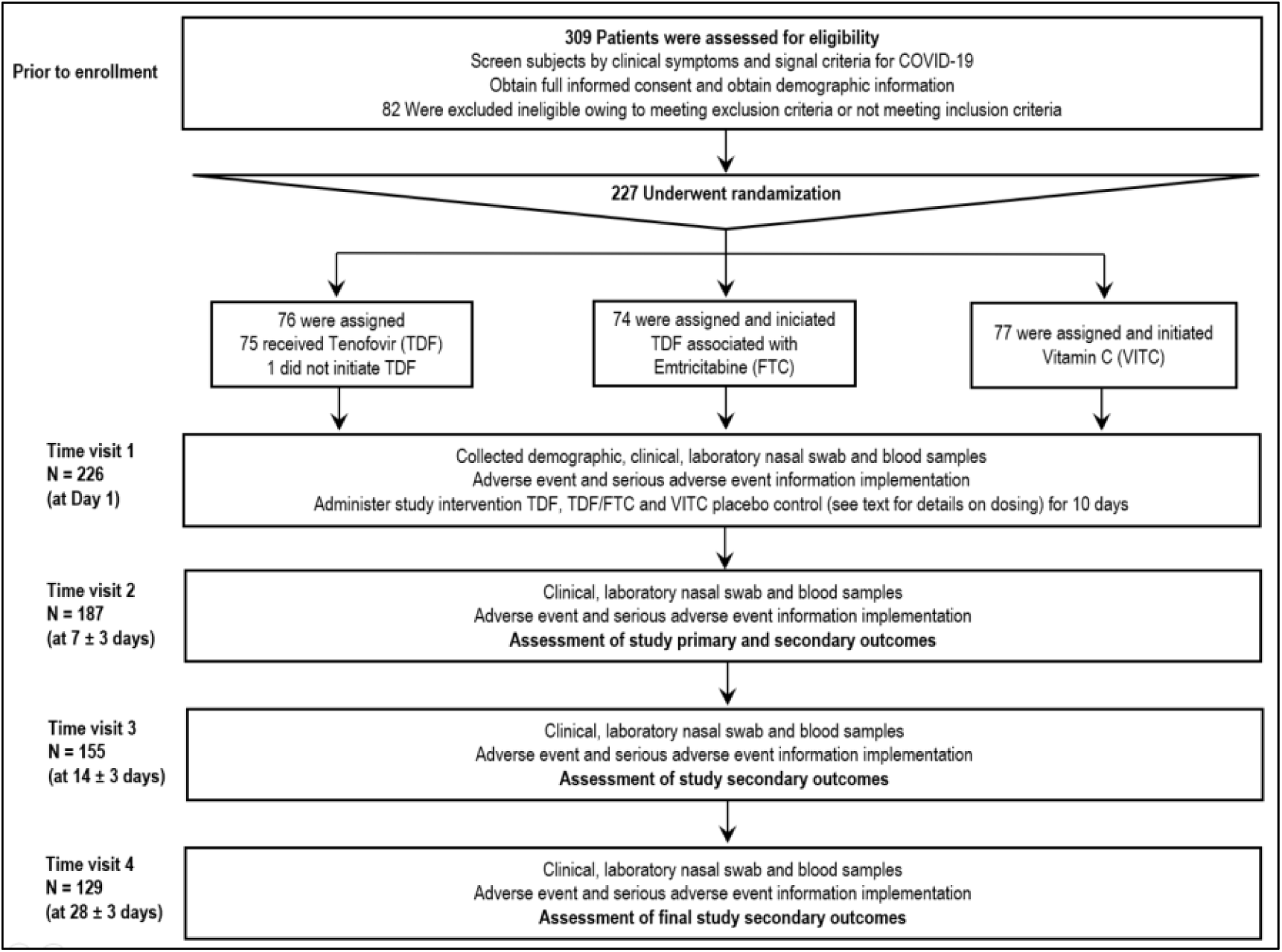
Enrollment, randomization, and study protocol follow up. Three hundred and nine patients with clinical suspicion of COVID-19 were selected and provided consent to record the demographic data and verify the inclusion and exclusion criteria for recruitment in the study. Eighty-two patients were excluded from the study for not meeting the inclusion criteria. Two hundred twenty-seven patients were randomized to receive one of three treatments: TDF (76 patients); TDF/FTC (74 patients) and Vitamin C placebo (77 patients). Only one patient in the TDF group did not start the medication, so 75 patients in this group started the treatment.

The clinical symptoms and signs in decreasing order of frequency were headache 83% (182/221), muscle pain 73% (158/217), rhinorrhea 71% (155/220), cough 69% (154/222), sore throat 68% (150/221), weakness 66% (145/221), fever (≥37.8°C) 53% (116/220), ageusia or dysgeusia 31% (68/220), anosmia or dysosmia 31% (67/220), shortness of breath 15% (32/218) and skin irritation 1% (1/218) (**Table 2**). Thirty-two percent (72/226) had more than one symptom or sign. Patients positive for SARS-CoV-2 showed significant differences for the following symptoms or signs: sore throat (62% vs 78%; p = 0.010; Chi-Square Test), fever (58% vs 43%; 0.031), ageusia or dysgeusia (36% vs 23%; 0.045), and anosmia or dysosmia (37% vs 19%; 0.005). As sore throat was more frequent in the negative group for SARS-CoV-2, the clinical symptoms and signs score to assess the effectiveness of drugs in the clinical trial was based only on the three most frequent clinical symptoms and signs associated with infections by SARS-CoV-2 (**Table 2**). **Table 3** summarizes the multivariate logistic analysis with the four symptoms and signs selected in the univariate analysis to determine predictive variables of SARS-CoV-2 infection. The logistic regression model showed that anosmia or dysosmia (Odds ratio = 2.70; p = 0.023) and sore throat (Odds ratio = 0.45; p = 0.019) were the two most significant symptoms and signs to distinguish infection from SARS-CoV-2. The model had an accuracy of 67%, sensitivity of 82%, specificity of 43%. The positive predictive value was 71% and the negative predictive value was 58%.

**Table 2.** Clinical characteristics and treatment of mild to moderate acute respiratory infections with SARS-CoV-2 infection and without SARS-CoV-2.

| Clinical signals and symptoms | All<br>N (%) | Patients with<br>Laboratory RT-PCR-Confirmed<br>SARS-CoV-2<br>N (%) | Patients who<br>Tested<br>Negative for<br>SARS-CoV-2<br>N (%) | P<br>values <sup>+</sup> |
| --- | --- | --- | --- | --- |
| Headache | 182 / 221 (83) | 113 / 139 (81) | 69 / 82 (84) | 0.591 |
| Muscle ache | 158 / 217 (73) | 100 / 136 (74) | 58 / 81 (72) | 0.758 |
| Rhinorrhea | 155 / 220 (71) | 96 / 138 (70) | 59 / 82 (72) | 0.708 |
| Cough | 154 / 222 (69) | 101 / 139 (73) | 53 / 83 (64) | 0.168 |
| Sore throat | 150 / 221 (68) | 85 / 138 (62) | 65 / 83 (78) | **0.010** |
| Weakness | 145 / 221 (66) | 86 / 138 (62) | 59 / 83 (71) | 0.184 |
| Fever ($\geq 37.8^{\circ}\text{C}$) | 116 / 220 (53) | 81 / 139 (58) | 35 / 81 (43) | **0.031** |
| Ageusia ou dysgeusia | 68 / 220 (31) | 49 / 137 (36) | 19 / 83 (23) | **0.045** |
| Anosmia ou disosmia | 67 / 220 (31) | 51 / 137 (37) | 16 / 83 (19) | **0.005** |
| Shortness of breath | 32 / 218 (15) | 22 / 137 (16) | 10 / 81 (12) | 0.454 |
| Rash | 1 / 218 (1) | 1 / 136 (1) | 0 / 82 (0) | 0.436 |
| More than one sign or symptom | 72 / 226 (32) | 54 / 139 (39) | 18 / 83 (22) | **0.008** |
| **Treatment <sup>§</sup>** | **N = 222** | **N = 139** | **N = 83** |  |
| Analgesic and antipyretic | 66 (30) | 16 (12) | 50 (60) | **<0.0001** |
| Antibiotic | 20 (9) | 4 (3) | 16 (19) | **0.0001** |
| Anti-inflammatory | 19 (9) | 4 (3) | 15 (18) | **0.0002** |
| Antihistamine | 16 (7) | 8 (6) | 8 (10) | 0.294 |
| Vitamins | 10 (5) | 4 (3) | 6 (7) | 0.181 |
| Ivermectin | 5 (2) | 1 (1) | 4 (5) | 0.066 |
| Antidepressant | 4 (2) | 3 (2) | 1 (1) | 1.000 |
| Nasal steroids | 4 (2) | - | 4 (5) | - |
| Syrup | 4 (2) | 1 (1) | 3 (4) | 0.148 |

<sup>+</sup> Statistical tests used: Chi-Square test Likelihood Ratio.
<sup>§</sup> Treatment used in three or more patients in the total.

**Table 3.**
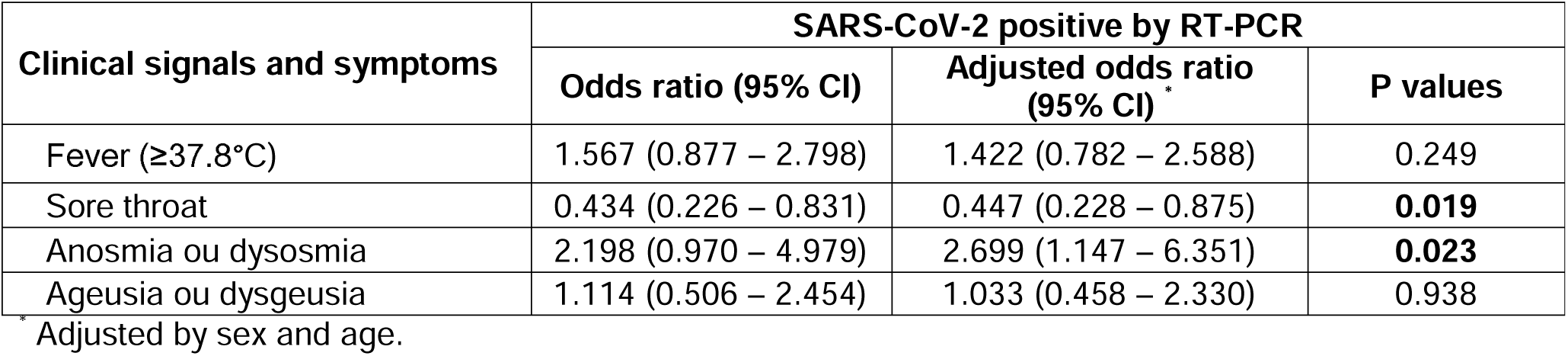
Multivariate logistic regression predictors of SARS-CoV-2 infections based in clinical signals and symptoms.

| Clinical signals and symptoms | SARS-CoV-2 positive by RT-PCR |  |  |
| --- | --- | --- | --- |
|  | Odds ratio (95% CI) | Adjusted odds ratio (95% CI) * | P values |
| Fever ($\geq 37.8^{\circ}\text{C}$) | 1.567 (0.877 – 2.798) | 1.422 (0.782 – 2.588) | 0.249 |
| Sore throat | 0.434 (0.226 – 0.831) | 0.447 (0.228 – 0.875) | **0.019** |
| Anosmia ou dysosmia | 2.198 (0.970 – 4.979) | 2.699 (1.147 – 6.351) | **0.023** |
| Ageusia ou dysgeusia | 1.114 (0.506 – 2.454) | 1.033 (0.458 – 2.330) | 0.938 |

\* Adjusted by sex and age.

The main drugs used concomitantly with the drugs in the clinical trial (**Table 2**) were in descending order: analgesics and antipyretics 30% (66/222), antibiotics 9% (20/222), anti-inflammatory drugs 9% (19/222), antihistamines 7% (16/222), vitamins 5% (10/222), and 2% for ivermectin (5/222), antidepressants (4/222), nasal steroids (4/222) and syrups (4 /222). Patients in the SARS-CoV-2 negative group showed higher proportions taking analgesics and antipyretics, antibiotics, and anti-inflammatory drugs (p < 0.05; Chi-Square Test). **Table 4** summarizes the variables for two clinical scores based on the four significant predictive symptoms and signs of mild to moderate respiratory infection associated with SARS-CoV-2. In the evaluation of the first score, on the second visit (7 ± 3 days), patients with no symptoms or signs had a proportion of 60% (73/122); one symptom or sign 11% (13/122), two symptoms or signs 28% (34/122); and three or more symptoms and signs 2% (2/122). In the second evaluated score, patients with none or one symptom or sign 71% (86/122) and two or more symptoms or signs 30% (36/122). When comparing the experimental groups of drugs used, there were no significant differences in the two clinical scores evaluated.

**Table 4.** Outcomes according to clinical scores on the ordinal scale treated population at 7^th^ day visit with COVID-19.

| Characteristic | All<br>(N = 122) | Tenofovir<br>(N = 43) | Tenofovir plus<br>Emtricitabine<br>(N = 38) | Placebo<br>Vitamin C<br>(N = 41) |
| --- | --- | --- | --- | --- |
| 1. Score on ordinal scale – no. (%) * |  |  |  |  |
| Positive qPCR for SARS-CoV-2 §: |  |  |  |  |
| 1. No symptom or signal | 73 (60) | 27 (63) <sup>a+</sup> | 19 (50) <sup>a</sup> | 27 (66) <sup>a</sup> |
| 2. One symptom or signal | 13 (11) | 4 (9) <sup>a</sup> | 4 (11) <sup>a</sup> | 5 (12) <sup>a</sup> |
| 3. Two symptom or signal | 34 (28) | 11 (26) <sup>a</sup> | 15 (40) <sup>a</sup> | 8 (20) <sup>a</sup> |
| 4. Three or more symptom or signal | 2 (2) | 1 (2) <sup>a</sup> | - | 1 (2) <sup>a</sup> |
| 2. Score of symptom or signal– no. (%) |  |  |  |  |
| Positive qPCR for SARS-CoV-2: |  |  |  |  |
| 1. No or one symptom or signal | 86 (71) | 31 (72) <sup>a</sup> | 23 (61) <sup>a</sup> | 32 (78) <sup>a</sup> |
| 2. Two or more symptom or signal | 36 (30) | 12 (28) <sup>a</sup> | 15 (40) <sup>a</sup> | 9 (22) <sup>a</sup> |

\* Percentage may not total 100 because of rounding. § qPCR positive by CT < 37.
<sup>+</sup> Each subscript letter denotes a subset of Grupo categories whose column proportions do not differ significantly from each other at the ,05 level. Statistical tests used: Chi-Square test Likelihood Ratio.

Adverse events and serious adverse events are shown in **Table 5**. Patients with mild to moderate respiratory infection, positive for SARS-CoV-2 and taking the drugs in the clinical trial presented the following adverse events in descending order: nausea 18% (18/122); diarrhea 7% (9/122); vomiting 5% (6/122); stomach pain 3% (4/122) and 2% (2/122) for abdominal pain, headache and insomnia. The comparison between the experimental groups did not show significant differences (p < 0.05). Five serious adverse events were recorded (4 hospitalizations for COVID-19 and 1 hospitalization for a firearm accident), and no deaths were reported following these recorded serious adverse events. None of these health problems were considered by the researchers to be related to the study medications.

**Table 5.**
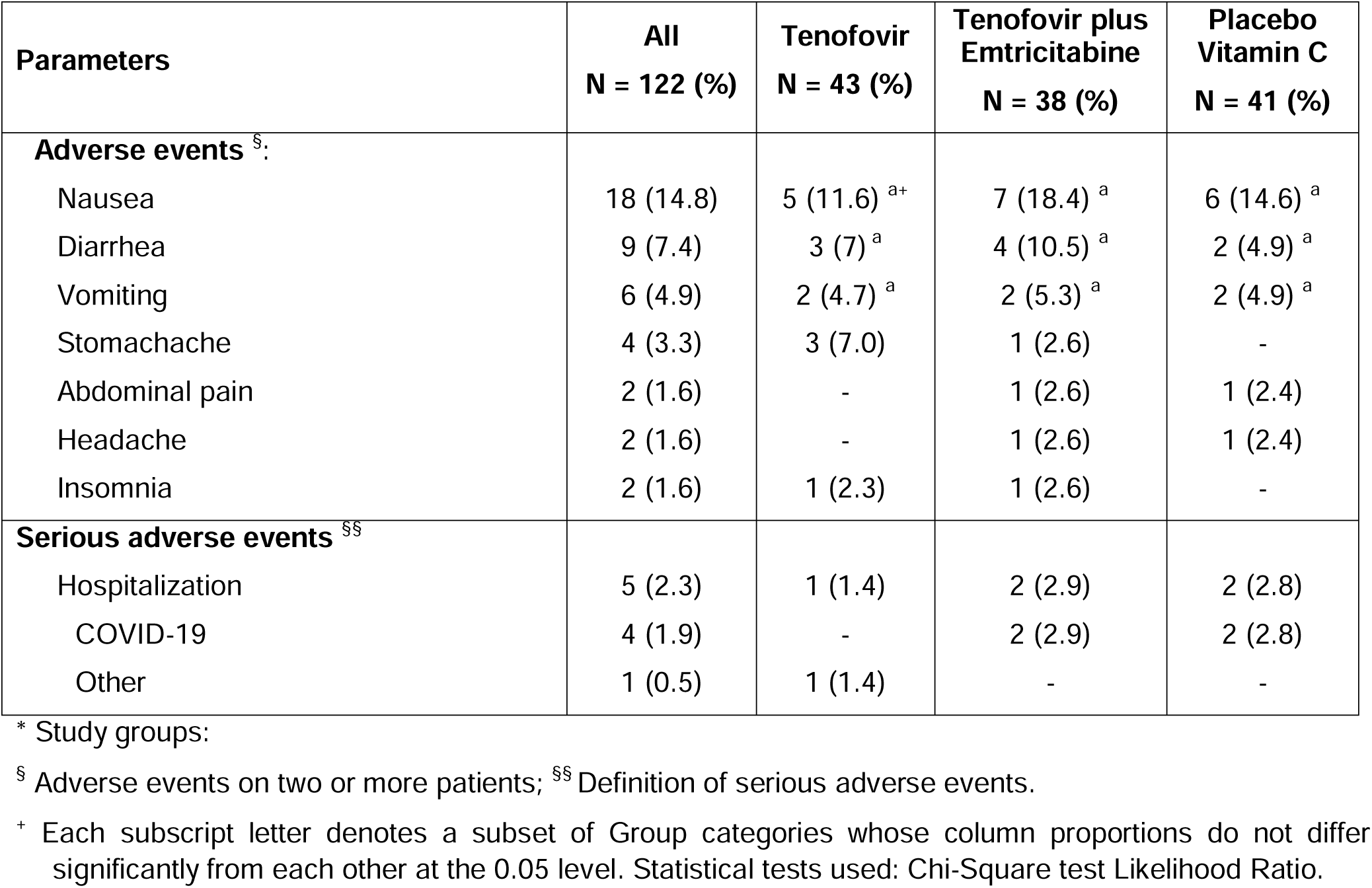
Adverse events and serious adverse events by overall and study groups* in treated patients with mild to moderate respiratory SARS-CoV-2 infection at 7^th^ day visit.

**Supplemental Tables S1, S2** and **S3** (**Appendix**) summarize data on protein biomarkers of pro and anti-inflammatory activity and cell growth factors IL-6, MCP-3, IL-1β, IL1-RA, IL-10, G-CSF, TNF-α, MCP-1, IFN-γ, and IP-10 in serum samples from patients with mild to moderate respiratory infection with or without SARS-CoV-2 in the first, third (14 ± 3 days) and fourth (28 ± 3 days) follow-up visits of patients. **Figure 3** shows the biomarkers with significant differences between patients with mild to moderate positive and negative respiratory infection for SARS-CoV-2. Levels of G-CSF, IL-1β, IL-6 and TNF-α biomarkers are significantly higher in patients with SARS-CoV-2 compared to patients negative for SARS-CoV-2 (p < 0.05; Mann -Whitney U Test). In the follow-up of biomarkers on the 14th. and 28th. days, the values of the biomarkers IL-10 and TNF-α, as well as IL-1β, IL-10 and MCP-3 remained higher respectively in the times observed in patients with SARS-CoV-2 infection compared to patients without infection by SARS-CoV-2 (p < 0.05; Mann-Whitney U Test). **Supplemental Table S4** summarizes the IgM and IgG immunoglobulin titer data measured at the 14^th^ visits. and 28^th^ days of follow-up of patients positive for SARS-CoV-2. IgM values did not change, but IgG values increased significantly by the 28^th^ day in relation to the 14^th^ day (p < 0.05; Wilcoxon Signed Rank Test paired samples) in patients in all treatment groups. Between the experimental groups of the clinical trial there were no significant differences in immunoglobulins at the times evaluated.

**Figure 3.**
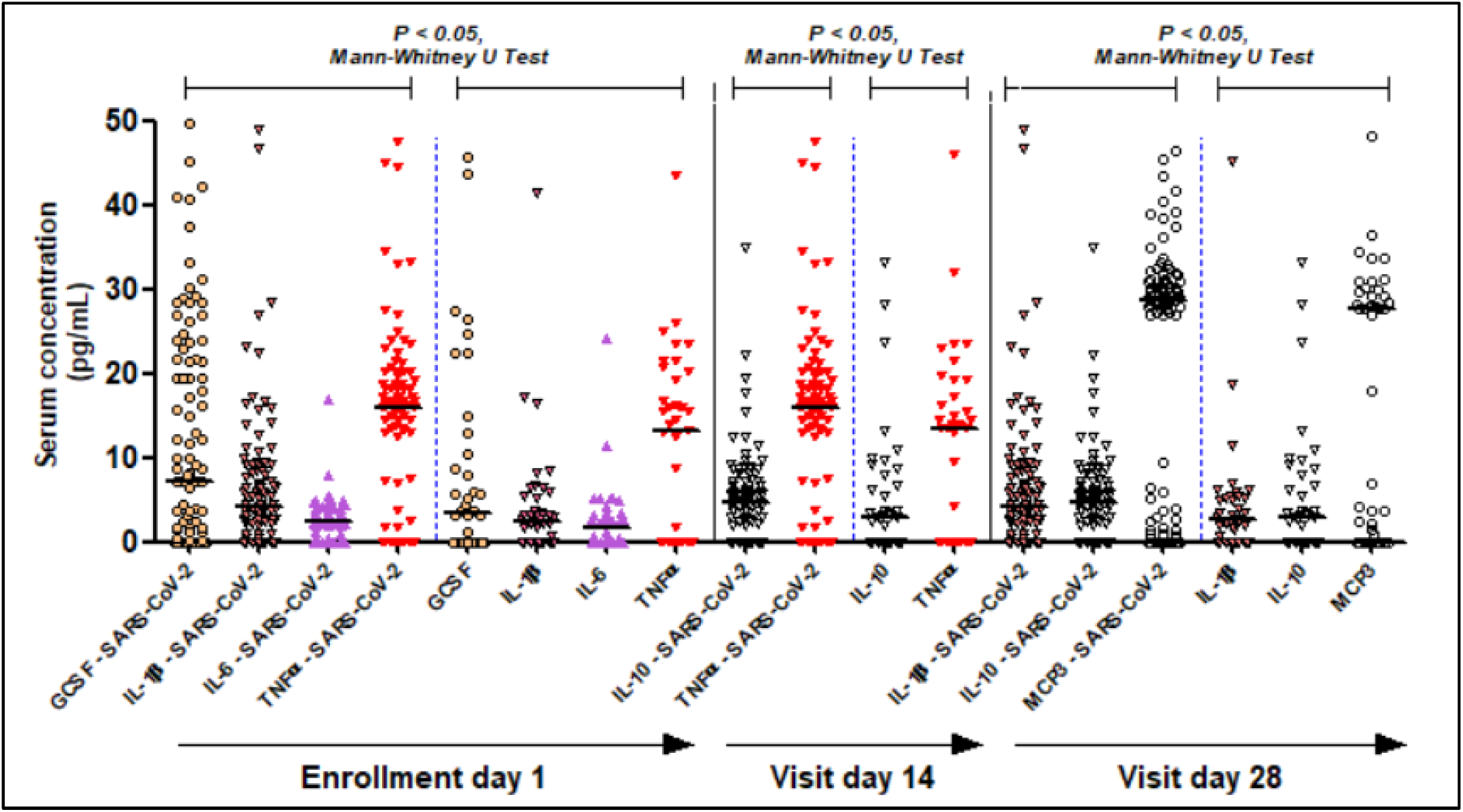
Cytokines, chemokines, and cell growth factor in systemic blood of patients with and without mild to moderate SARS-CoV-2 infection. This figure shows the biomarkers with significant differences between patients with mild to moderate positive and negative respiratory infection for SARS-CoV-2. Levels of G-CSF, IL-1β, IL-6 and TNF-α biomarkers are significantly higher in patients with SARS-CoV-2 compared to patients negative for SARS-CoV-2 (p < 0.05; Mann -Whitney U Test). In the follow-up of biomarkers on the 14th. and 28th. days, the values of the biomarkers IL-10 and TNF-α, as well as IL-1β, IL-10 and MCP-3 remained higher respectively in the times observed in patients with SARS-CoV-2 infection compared to patients without infection by SARS-CoV-2 (p < 0.05; Mann-Whitney U Test).

## Discussion

Mild to moderate acute respiratory illnesses are considered the leading causes of visits for outpatients of all ages^13^. This is largely caused by viruses and in the pandemic phase of COVID-19 makes accurate diagnosis at the clinical and laboratory level necessary, and is thus of economic, socio-sanitary and in defined parameters for clinical trial research and in public health. The analysis at baseline of study recruitment (**Table 2**) determined whether there were signs and symptoms that could help physicians discriminate COVID-19 from other situations suggestive of mild to moderate respiratory infection in the absence of SARS-CoV-2 infection. The main symptoms most associated in patients with COVID-19 in this prospective clinical trial study were fever (≥37.8°C), ageusia or dysgeusia, and anosmia or dysosmia. Sore throat symptom was associated with mild to moderate respiratory infection in the absence of SARS-CoV-2 infection. Multivariate logistic analysis of these symptoms adjusted for gender and age of patients showed the persistence of two symptoms, sore throat, and anosmia or dysosmia. In this model, accuracy was 67% with sensitivity in efficacy and specificity of 82% and 43%, respectively, positive predictive value of 71% compared to 62% reliability of “practitioner’s clinical intuition” as described, and negative predictive of 58%. Studies before coronavirus epidemics showed consistency with this model and to determine signs and symptoms more associated with influenza infection showed that fever (feeling of fever or chills), cough, myalgia and weakness were more frequent and, in several studies, showed the predictive value positive for cough and fever^14,15^. A recent study from the group before the pandemic showed that severe acute respiratory infections in the absence of SARS-CoV-2 infection in the same population and geographic region studied were mainly associated with influenza A (H1N1) and influenza B viruses^16^. Thus, during the period of the COVID-19 epidemic when SARS-CoV-2 is more present in the community, a high index of suspicion of the disease is guaranteed in patients who present with an acute onset of fever (≥37.8°C), ageusia or dysgeusia and anosmia or dysosmia. Two scores based on these symptoms (**Table 4**) were delineated as primary parameters and most associated with SARS-CoV-2 infection to assess efficacy of TDF and TDF combined with FTC in patients on the seventh day of COVID-19 evolution.

### Mild to Moderate COVID-19 Biomarkers

This study focuses on patients with suspected mild to moderate SARS-CoV-2 respiratory infection. In this sense, the pathobiology of SARS-CoV-2 infection predominates in the upper airways, the predominant site of expression in the host of angiotensin-2 converting enzyme (ACE2) receptors^17^. The S protein of SARS-CoV-2 binds to the ACE2 receptor on the pulmonary epithelial cell, allowing the virus to fuse with the host cell and supported by the action of dipeptidyl peptidase 4 (DPP4; or CD26)^17,18^. The S protein of SARS-CoV-2 binds with the ACE2 receptor 10-20 times more potent than the SARS-CoV virus^18^. ACE2 is widely expressed in lung epithelial cells and other cells such as macrophages in lung tissue^19,20^ and has little or no expression in human peripheral blood immune system cells^20^. Recent evidence has shown that the accessory protein encoded by the ORF7a of SARS-CoV-2 is responsible for activating the transcription factor NF-kB associated with the expression of pro-inflammatory cytokines including IL-1alpha, IL-1beta, IL-6, IL-8, IL-10, TNF-alpha and IFNbeta^21^. The present work shows a profile of pro-inflammatory cytokines such as IL-1beta, IL-6 and TNF-alpha in addition to G-CSF increased in mild to moderate respiratory infections caused by SARS-CoV-2 when compared to other respiratory infections caused by seasonal respiratory viruses, suggesting a profile of cytokines produced by activation of lung tissue macrophages. Recent work comparing the cytokine profile in patients with severe COVID-19 and severe influenza has shown a similar increase in pro-inflammatory cytokines in these patients, showing similarity in pathobiology even when observed in cases of severe COVID-19^22^. **Figure 4** summarizes the model designed of these findings for the pathobiology of mild to moderate respiratory infection by SARS-CoV-2.

**Figure 4.**
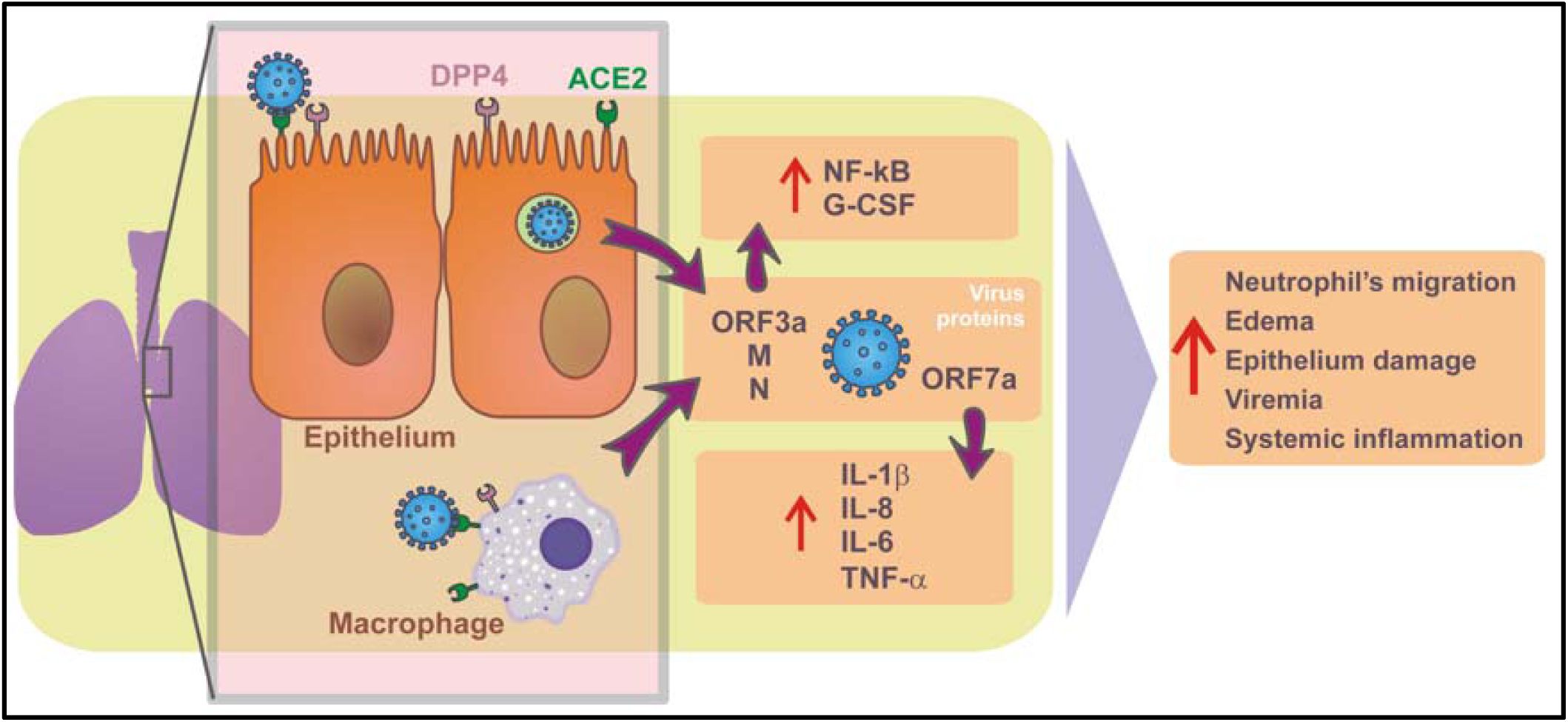
Model of the pathobiology of mild to moderate respiratory SARS-CoV-2 human infection. The pathobiology of SARS-CoV-2 infection predominates in the upper airways in these patients enrolled in the study protocol, the predominant site of expression in the host of angiotensin-2 converting enzyme (ACE2) receptors^17^. The S protein of SARS-CoV-2 binds to the ACE2 receptor on the pulmonary epithelial cell, allowing the virus to fuse with the host cell and supported by the action of dipeptidyl peptidase 4 (DPP4; or CD26)^17,18^. ACE2 is widely expressed in lung epithelial cells and other cells such as macrophages in lung tissue^19,20^ and has little or no expression in human peripheral blood immune system cells^20^. Recent evidence has shown that the accessory protein encoded by the ORF7a of SARS-CoV-2 is responsible for activating the transcription factor NF-kB associated with the expression of pro-inflammatory cytokines including IL-1alpha, IL-1beta, IL-6, IL-8, IL-10, TNF-alpha and IFNbeta^21^. The present work shows a profile of pro-inflammatory cytokines such as IL-1beta, IL-6 and TNF-alpha in addition to G-CSF increased in mild to moderate respiratory infections caused by SARS-CoV-2 when compared to other respiratory infections caused by seasonal respiratory viruses, suggesting a profile of cytokines produced by activation of lung tissue macrophages.

### Efficacy and Toxicity of TDF and TDF Combined with FTC in COVID-19

Antiviral therapies alone or in combination are important measures for the prevention and treatment of emerging viruses such as SARS-CoV-2. Yet several vaccines are effective in preventing COVID-19 in various parts of the world^23-25^, there is still no proven effective antiviral treatment for COVID-19. The RNA-dependent RNA polymerase (RdRp) of coronaviruses is an interesting target for antivirals^26^. Among the various drugs tested, remdesivir (adenosine analogue) was initially tested during the COVID-19 pandemic because of its high potency of inhibition for RdRp^4,27^ and wide action against several viruses of RNA including Ebola virus, hepatitis C, SARS-CoV and MERS-CoV^4,27^. Remdesivir was superior to placebo in reducing recovery time in adults hospitalized with COVID-19 and evidence of lower respiratory tract infection. On October 22, 2020, the US Food and Drug Administration approved the first antiviral drug called Veklury (remdesivir) for the treatment of COVID-19 that requires hospitalization in patients over 12 years of age^28^. TDF, which has a similar structure to remdesivir, competitively inhibits the RdRp of HIV and hepatitis B viruses, being well tolerated and preventing the progression of liver fibrosis^28-36^, has an inhibitory effect on the RdRp of SARS-CoV-2^6,37^. TDF is a low-cost, widely available generic drug with low toxicity recommended in combination with FTC (cytosine analogue) and/or lamivudine (3TC; cytosine analogue) for effective combined antiretroviral therapy^29-32^. Considering these data and the existence of a drug already licensed containing an association of TDF with FTC, we decided to carry out the present clinical study controlled with a placebo, including groups treated with TDF alone and with TDF combined with FTC. Several reports of preclinical studies of these drugs for SARS-CoV-2 have been found, including several in silico studies, which have shown the ability of TDF to bind to RdRp of SARS-CoV-2^7,38-46^. TDF also binds with other important virus proteins, including the papain-like protease (PLpro) and the main or 3C-like protease (Mpro/3CLpro)^47-49^, in addition to interacting with the S protein and the ACE2 receptor^50,51^, suggesting multiple binding and interference targets in the replication cycle of the virus. *In vitro* studies have shown contradictory results in the efficacy of TDF, but some have shown efficacy in reducing viral load in cell cultures and viral titers in nasopharyngeal samples in the experimental SARS-CoV-2 infection model in ferrets^6-8^. Some clinical studies suggest that TDF combined with antiretroviral therapy in HIV-infected patients has reduced the incidence of COVID-19, viral load, risk of hospitalization, and death^52-54^. These clinical data are few and do not provide conclusive evidence suggesting the need for further clinical trials to determine the efficacy of TDF and TDF/ FTC in COVID-19. In the present study TDF and TDF/ FTC were evaluated in patients with mild to moderate COVID-19 and revealed that the groups treated with TDF and TDF / FTC did not differ from each other, nor in comparison with the group treated with placebo. As the selected patients had mild to moderate COVID-19, the viral load on the seventh day of evolution significantly decreased in all experimental groups, not allowing the assessment of drugs in this parameter. The documented adverse events and serious adverse events were of low frequency and there were no differences between the experimental groups in the clinical trial, showing consistency with the literature regarding the low clinical toxicity of TDF and TDF combined with FTC.

The use of viral load and signs and symptoms scores on the seventh day of illness were a limitation of the present study. As noted, the viral load is practically zero on the seventh day of illness in all experimental groups, which makes it difficult to evaluate the drugs in their antiviral action. As the selected COVID-19 cases had mild to moderate disease, it is likely that the majority had already recovered regardless of the drugs. Considering that most patients included in the study made adequate use of the drugs, judging by the control of their intake, we might have observed differences if we had followed symptoms and signs and viral load earlier, before the seventh day of illness.

### Summary of conclusions

The presence of fever (≥37.8°C), anosmia or dysosmia, ageusia or dysgeusia, with two or more symptoms in patients with mild to moderate respiratory infection, indicate the diagnosis of COVID-19. The score based on these symptoms and signs was useful for use in the clinical trial and for the implementation of preventive measures in the transmission of COVID-19. Univariate and multivariate logistic regression analysis for the predictive parameters of symptoms and signs associated with mild to moderate COVID-19 showed that anosmia or dysosmia symptoms, in the absence of sore throat, have accuracy and high sensitivity to predict COVID-19, compared to mild and moderate respiratory infections due to causes other than SARS-CoV-2. Pharmacological intervention with TDF and TDF / FTC did not change clinical signs and symptoms scores in mild to moderate SARS-CoV-2 respiratory infection. Likewise, it did not change the seroconversion to IgM and IgG, nor biomarkers of pro and anti-inflammatory activity. Patients with mild to moderate respiratory infections positive for SARS-CoV-2 had a pro-inflammatory systemic profile with increased biomarkers G-CSF, IL-1β, IL-6 and TNF-α when compared with patients negative for SARS-CoV-two. In addition, the profile of IL-10 and TNF-α biomarkers, as well as IL-1β, IL-10 and MCP-3 remained at higher concentrations in patients with SARS-CoV-2 infection compared to patients without infection by SARS-CoV-2.

This article was preprinted at https://www.medrxiv.org/content/…

## Data Availability

ClinicalTrials.gov Identifier: NCT04712357

## Acknowledgments

We thank the staff and patients at Hospital São José, SESA, CE and the Núcleo de Biomedicina, Faculdade de Medicina, Universidade Federal do Ceará for their availability, dedication and work done for this study.

ARTAN-C19 proposal was supported by Rede Vírus, Ministério da Ciência, Tecnologia, Inovações e Comunicações (MCTIC), Brasília-DF through CNPq, Process number: 403542/2020-0.

## Author Contributions

Conceptualization, A.A. Santos, P.J.C. Magalhães, A. Havt, N.P. Lopes, E. Arruda-Neto, A.A.M. Lima; methodology, E.A.G. Arruda, R.J. Pires-Neto, M.S. Medeiros, J. Quirino-Filho, M. Clementino, R.N.D.G. Gondim, L.M.V.C. Magalhães, K.F. Cavalcante, V.A.F. Viana, Liana Perdigão Mello, R.B Martins, A. Havt, N.P. Lopes, E. Arruda-Neto, A.A.M. Lima; formal analysis, E.A.G. Arruda, J. Quirino-Filho, M. Clementino, R.N.D.G. Gondim, A. Havt, E. Arruda-Neto, A.A.M. Lima; data curation, E.A.G. Arruda, R.J. Pires-Neto, M.S. Medeiros, J. Quirino-Filho, M. Clementino, R.N.D.G. Gondim, L.M.V.C. Magalhães, K.F. Cavalcante, V.A.F. Viana, Liana Perdigão Mello, R.B Martins, A. Havt, A.A.M. Lima; writing-original draft preparation, E.A.G. Arruda, M. Clementino, R.N.D.G. Gondim, L.M.V.C. Magalhães, A.A. Santos, P.J.C. Magalhães, A. Havt, E. Arruda-Neto, A.A.M. Lima; writing-review and editing, E.A.G. Arruda, M. Clementino, R.N.D.G. Gondim, L.M.V.C. Magalhães, A.A. Santos, P.J.C. Magalhães, A. Havt, N.P. Lopes, E. Arruda-Neto, A.A.M. Lima. All authors have read and agreed to the published version of the manuscript.

## Supplement Tables

**Table S1 Supplement.** Human cytokines, chemokines and growth factor serum concentration, pg/mL, in patients with mild to moderate respiratory infection with and without SARS-CoV-2 at enrollment.

| At enrollment Day 1 | | G-CSF<sup>#</sup> | IFN-γ | IL-1β<sup>*</sup> | IL-1RA | IL-6<sup>*</sup> | IL-10 | IP-10 | MCP-1 | MCP-3 | TNF-α<sup>*</sup> |
| --- | --- | --- | --- | --- | --- | --- | --- | --- | --- | --- | --- |
| **SARS-CoV-2 infection (N=139)** | Median | 10 | 23 | 5 | 5 | 3 | 5 | 123 | 328 | 28 | 16 |
|  | Interquartile range | 0-29 | 0-108 | 1-10 | 0-7 | 0-4 | 0-9 | 39-297 | 105-690 | 2-32 | 1-23 |
| **Non-SARS-CoV-2 infection (N=83)** | Median | 3 | 8 | 3 | 4 | 2 | 2 | 53 | 184 | 19 | 13 |
|  | Interquartile range | 0-14 | 0-44 | 0-5 | 0-5 | 0-3 | 0-7 | 21-234 | 74-448 | 0-31 | 0-20 |

<sup>#</sup> G-CSF: granulocyte-colony stimulating factor; IFN-γ : interferon-gamma; IL-1β : interleukin-1 beta; IL-1RA: interleukin 1 receptor antagonist; IL-6: interleukin 6; IL-10: interleukin 10; IP-10: interferon $\gamma$ -induced protein 10kDa or C-X-C motif chemokine 10 (CXC10); MCP-1: monocyte chemoattractant protein-1; MCP-3: human monocyte chemotactic protein-3 or chemokine (C-C motif) ligand 7 (CCL7); and TNF-α : tumor necrosis factor alpha. <sup>\*</sup> P < 0.05 SARS-CoV-2 versus non-SARS-CoV-2 infection (Mann-Whitney U Test).

**Table S2 Supplement.** Human cytokines, chemokines and growth factor serum concentration, pg/mL, in patients with mild to moderate respiratory infection with and without SARS-CoV-2 at visit day 14^th^.

| Visit Day 14<sup>o</sup>. | | G-CSF<sup>#</sup> | IFN-γ | IL-1β | IL-1RA | IL-6 | IL-10<sup>*</sup> | IP-10 | MCP-1 | MCP-3 | TNF-α<sup>*</sup> |
| --- | --- | --- | --- | --- | --- | --- | --- | --- | --- | --- | --- |
| **SARS-CoV-2 infection (N=139)** | Median | 7 | 9 | 4 | 4 | 2 | 5 | 87 | 293 | 29 | 16 |
|  | Interquartile range | 0-23 | 0-63 | 2-9 | 0-7 | 0-3 | 0-7 | 31-261 | 122-772 | 2-31 | 0-20 |
| **Non-SARS-CoV-2 infection (N=83)** | Median | 2 | 1 | 3 | 4 | 2 | 3 | 78 | 212 | 28 | 13 |
|  | Interquartile range | 0-16 | 0-59 | 0-5 | 0-6 | 0-3 | 0-8 | 22-278 | 92-606 | 1-30 | 0-19 |

<sup>#</sup> G-CSF: granulocyte-colony stimulating factor; IFN-γ : interferon-gamma; IL-1β : interleukin-1 beta; IL-1RA: interleukin 1 receptor antagonist; IL-6: interleukin 6; IL-10: interleukin 10; IP-10: interferon $\gamma$ -induced protein 10kDa or C-X-C motif chemokine 10 (CXC10); MCP-1: monocyte chemoattractant protein-1; MCP-3: human monocyte chemotactic protein-3 or chemokine (C-C motif) ligand 7 (CCL7); and TNF-α : tumor necrosis factor alpha. <sup>\*</sup> P < 0.05 SARS-CoV-2 versus non-SARS-CoV-2 infection (Mann-Whitney U Test).

**Table S3 Supplement.** Human cytokines, chemokines and growth factor serum concentration, pg/mL, in patients with mild to moderate respiratory infection with and without SARS-CoV-2 at visit day 28^th^.

| Visit Day 28<sup>o</sup>. | | G-CSF<sup>#</sup> | IFN-γ | IL-1β<sup>*</sup> | IL-1RA | IL-6 | IL-10<sup>*</sup> | IP-10 | MCP-1 | MCP-3<sup>*</sup> | TNF-α |
| --- | --- | --- | --- | --- | --- | --- | --- | --- | --- | --- | --- |
| **SARS-CoV-2 infection (N=139)** | Median | 6 | 13 | 5 | 4 | 2 | 4 | 102 | 403 | 29 | 16 |
|  | Interquartile range | 0-23 | 0-51 | 2-9 | 0-7 | 0-3 | 0-7 | 30-225 | 120-790 | 3-31 | 0-22 |
| **Non-SARS-CoV-2 infection (N=83)** | Median | 0 | 4 | 2 | 4 | 2 | 2 | 57 | 210 | 27 | 13 |
|  | Interquartile range | 0-15 | 0-51 | 0-7 | 0-5 | 0-3 | 0-7 | 23-232 | 90-528 | 2-30 | 0-19 |

<sup>#</sup> G-CSF: granulocyte-colony stimulating factor; IFN-γ : interferon-gamma; IL-1β : interleukin-1 beta; IL-1RA: interleukin 1 receptor antagonist; IL-6: interleukin 6; IL-10: interleukin 10; IP-10: interferon $\gamma$ -induced protein 10kDa or C-X-C motif chemokine 10 (CXC10); MCP-1: monocyte chemoattractant protein-1; MCP-3: human monocyte chemotactic protein-3 or chemokine (C-C motif) ligand 7 (CCL7); and TNF-α : tumor necrosis factor alpha. <sup>\*</sup> P < 0.05 SARS-CoV-2 versus non-SARS-CoV-2 infection (Mann-Whitney U Test).

**Table S4 Supplement.**
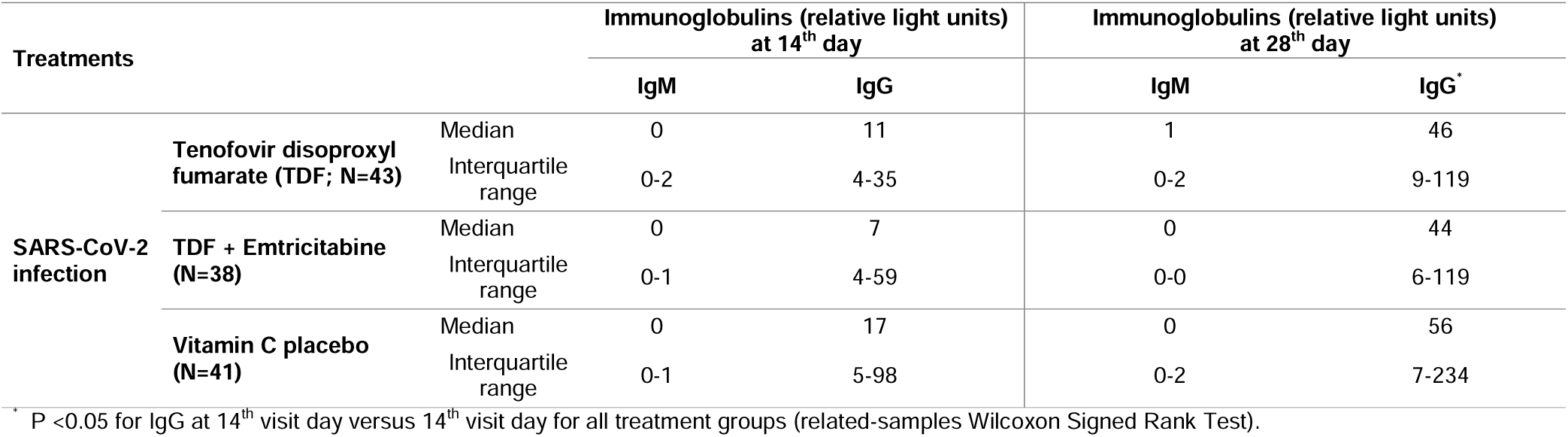
Immunoglobulins IgM and IgG serum concentrations in patients with mild to moderate respiratory SARS-CoV-2 infection.

**Members of the Antiretroviral Nucleotide Analogs Study Group in COVID-C19 Study Group - ARTAN-C19.**

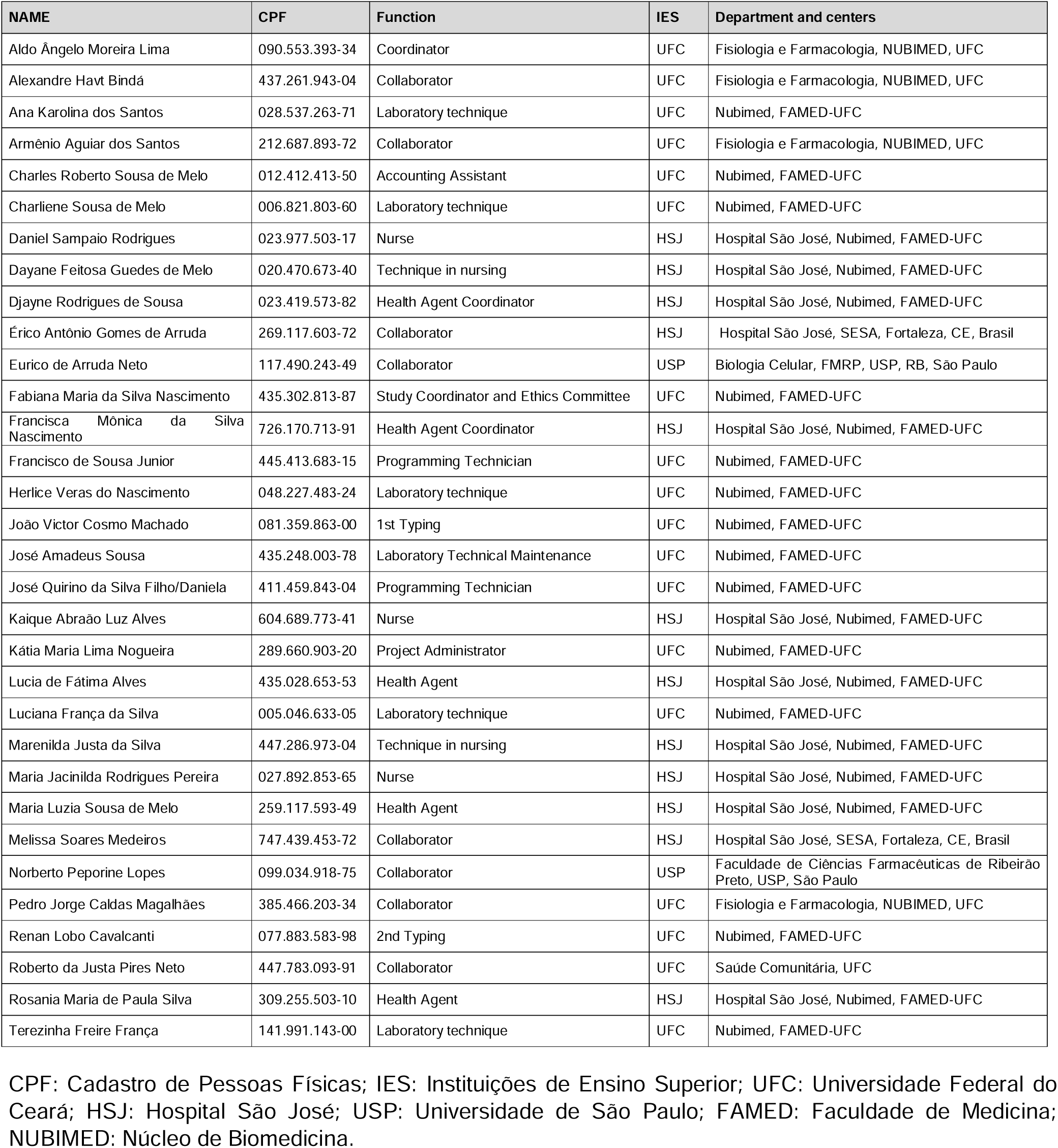

## References

1. Pormohammad A, Ghorbani S, Khatami A, et al. Comparison of influenza type A and B with COVID-19: A global systematic review and meta-analysis on clinical, laboratory and radiographic findings. Rev Med Virol. 2020;e2179.

2. Maria Rosaria Barillari, Luca Bastiani, Jerome R Lechien, et al. A structural equation model to examine the clinical features of mild-to-moderate COVID-19: A multicenter Italian study. J Med Virol. 2021 Feb;93(2):983–994.

3. Mihaja Raberahona, Rado Rakotomalala, Etienne Rakotomijoro, et al. Clinical and epidemiological features discriminating confirmed COVID-19 patients from SARS-CoV-2 negative patients at screening centres in Madagascar. Int J Infect Dis. 2021;103:6–8.

4. J.H. Beigel, K.M. Tomashek, L.E. Dodd, et al., for the ACTT-1 Study Group Members.Remdesivir for the Treatment of Covid-19 — Final Report. N Engl J Med. 2020 Nov 5;383(19):1813–1826.

5. Zanella I, Zizioli D, Castelli F, Quiros-Roldan E. Tenofovir, Another Inexpensive, Well-Known and Widely Available Old Drug Repurposed for SARS-COV-2 Infection. Pharmaceuticals (Basel). 2021 May 11;14(5):454.

6. Giuliano C Clososki, Soldi RA, Silva RM, et al. Tenofovir disoproxil fumarate: new chemical developments and encouraging in vitro biological results for SARS-CoV-2. J Braz Chem Soc 2020 June.

7. Abdo A Elfiky. SARS-CoV-2 RNA Dependent RNA Polymerase (RdRp) Targeting: An in silico. Perspective. J Biomol Struct Dyn. 2021 Jun;39(9):3204–3212.

8. Su-Jin Park, Kwang-Min Yu, Young-Il Kin, Se-MI Kim et al. Antiviral Efficacies of FDA-Approved Drugs against SARS-CoV-2 Infection in Ferrets. mBio 2020; 11:e01114-20.

9. Weekly Report COVID-19, Year 2021, 32nd. Epidemiological Week, Municipal Health Department, Fortaleza-CE.

10. CDC-006-00019, Revision: 03; https://www.fda.gov/media/134922/download.

11. Good Clinical Practice Handbook. Division of Microbiology and Infectious Diseases, National Institute of Allergy and Infectious Diseases, National Institutes of Health, Department of Health and Human Services; Bethesda, MD, 2019.

12. Code of Federal Regulations & ICH Guidelines, 2019.

13. Uyeki TM. Influenza. Ann Intern Med. 2017 Sep 5;167(5):ITC33–ITC48.

14. Long CE, Hall CB, Cunningham CK, Weiner LB, Alger KP, Gouveia M, Colella CB, Schnabel KC, Barker WH. Influenza surveillance in community-dwelling elderly compared with children. Arch Fam Med. 1997;6(5):459–65.

15. AS Monto, S Gravenstein, M Elliott, M Colopy, J Schweinle. Clinical signs and symptoms predicting influenza infection. Arch Intern Med 2000;160(21):3243–7.

16. Filho JQ, Junior FS, Lima TBR, Viana VAF, Burgoa JSV, Soares AM, Leite ÁM, Herron SA, Newland HL, Sarnaik KS, Hanson GF, Papin JA, Moore SR, Lima AAM. Perinatal Outcomes of Asynchronous Influenza Vaccination, Ceará, Brazil, 2013-2018. Emerg Infect Dis. 2021;27(9):2409–2420.

17. Hou YJ, Okuda K, Edwards CE, Martinez DR, Asakura T, Dinnon KH 3rd, Kato T, Lee RE, Yount BL, Mascenik TM, Chen G, Olivier KN, Ghio A, Tse LV, Leist SR, Gralinski LE, Schäfer A, Dang H, Gilmore R, Nakano S, Sun L, Fulcher ML, Livraghi-Butrico A, Nicely NI, Cameron M, Cameron C, Kelvin DJ, de Silva A, Margolis DM, Markmann A, Bartelt L, Zumwalt R, Martinez FJ, Salvatore SP, Borczuk A, Tata PR, Sontake V, Kimple A, Jaspers I, O’Neal WK, Randell SH, Boucher RC, Baric RS. SARS-CoV-2 Reverse Genetics Reveals a Variable Infection Gradient in the Respiratory Tract. Cell. 2020;182(2):429-446.e14.

18. Wrapp D, Wang N, Corbett KS, et al. Cryo-EM structure of the 2019-nCoV spike in the prefusion conformation. Science. 2020;367:1260–1263.

19. Kuba K, Imai Y, Rao S, et al. A crucial role of angiotensin converting enzyme 2 (ACE2) in SARS coronavirus-induced lung injury. Nat Med. 2005;11:875–879.

20. Xiang Song, Wei Hu, Haibo Yu, Laura Zhao, Yeqian Zhao, Xin Zhao, Hai-Hui Xue, Yong Zhao. Little to no expression of angiotensin-converting enzyme-2 on most human peripheral blood immune cells but highly expressed on tissue macrophages. Cytometry A. 2020 Dec 6.

21. Su CM, Wang L, Yoo D. Activation of NF-κB and induction of proinflammatory cytokine expressions mediated by ORF7a protein of SARS-CoV-2. Sci Rep. 2021;11(1):13464.

22. Karaba AH, Zhou W, Hsieh LL, Figueroa A, Massaccesi G, Rothman RE, Fenstermacher KZJ, Sauer L, Shaw-Saliba K, Blair PW, Robinson ML, Leung S, Wesson R, Alachkar N, El-Diwany R, Ji H, Cox AL. Differential Cytokine Signatures of SARS-CoV-2 and Influenza Infection Highlight Key Differences in Pathobiology. Clin Infect Dis. 2021 May 20:ciab376.

23. Available online: https://www.who.int/news-room/q-a-detail/coronavirus-disease-(covid-19)-vaccines (accessed on 12 April 2021).

24. Kumar, A.; Dowling, W.E.; Román, R.G.; Chaudhari, A.; Gurry, C.; Le, T.T.; Tollefson, S.; Clark, C.E.; Bernasconi, V.; Kristiansen, P.A. Status Report on COVID-19 Vaccines Development. Curr Infect Dis Rep. 2021;23(6):9.

25. Golob, J.L.; Lugogo, N.; Lauring, A.S.; Lok, A.S. SARS-CoV-2 vaccines: A triumph of science and collaboration. JCI Insight. 2021;6(9):e149187.

26. Jeong, G.U.; Song, H.; Yoon, G.Y.; Kim, D.; Kwon, Y.-C. Therapeutic Strategies against COVID-19 and Structural Characterization of SARS-CoV-2: A Review. Front. Microbiol. 2020, 11, 1723.

27. Gordon CJ, Tchesnokov EP, Woolner E, Perry JK, Feng JY, Porter DP, Götte M. Remdesivir is a direct-acting antiviral that inhibits RNA-dependent RNA polymerase from severe acute respiratory syndrome coronavirus 2 with high potency. J Biol Chem. 2020;295(20):6785–6797.

28. https://www.fda.gov/news-events/press-announcements/fda-approves-first-treatment-covid-19.

29. Mu, Y.; Pham, M.; Podany, A.T.; Cory, T.J. Evaluating emtricitabine + rilpivirine + tenofovir alafenamide in combination for the treatment of HIV-infection. Expert Opin. Pharmacother. 2020, 21, 389–397.

30. Waters, L.; Mehta, V.; Gogtay, J.; Boffito, M. The evidence for using tenofovir disoproxil fumarate plus lamivudine as a nucleoside analogue backbone for the treatment of HIV. J. Virus Erad. 2021, 7, 100028.

31. Santevecchi, B.A.; Miller, S.; Childs-Kean, L.M. Doing More with Less: Review of Dolutegravir-Lamivudine, a Novel Single-Tablet Regimen for Antiretroviral-Naïve Adults with HIV-1 Infection. Ann. Pharmacother. 2020, 54, 1252–1259.

32. Buchbinder, S.P.; Liu, A.Y. CROI 2019: Advances in HIV prevention and plans to end the epidemic. Top. Antivir. Med. 2019, 27,8–25.

33. Marrazzo, J.M.; Rabe, L.; Kelly, C.; Richardson, B.; Deal, C.; Schwartz, J.L.; Chirenje, Z.M.; Piper, J.; Morrow, R.A.; Hendrix, C.W.; et al. Tenofovir Gel for Prevention of Herpes Simplex Virus Type 2 Acquisition: Findings from the VOICE Trial. J. Infect. Dis. 2019, 219, 1940–1947.

34. Chaix, M.-L.; Charreau, I.; Pintado, C.; Delaugerre, C.; Mahjoub, N.; Cotte, L.; Capitant, C.; Raffi, F.; Cua, E.; Pialoux, G.; et al. Effect of On-Demand Oral Pre-exposure Prophylaxis with Tenofovir/Emtricitabine on Herpes Simplex Virus-1/2 Incidence among Men Who Have Sex with Men: A Substudy of the ANRS IPERGAY Trial. Open Forum. Infect. Dis. 2018, 5.

35. Andrei, G.; Gillemot, S.; Topalis, D.; Snoeck, R. The Anti–Human Immunodeficiency Virus Drug Tenofovir, a Reverse Transcriptase Inhibitor, Also Targets the Herpes Simplex Virus DNA Polymerase. J. Infect. Dis. 2017, 217, 790–801.

36. Celum, C.; Hong, T.; Cent, A.; Donnell, D.; Morrow, R.; Baeten, J.M.; Firnhaber, C.; Grinsztejn, B.; Hosseinipour, M.C.; Lalloo, U.; et al. Herpes Simplex Virus Type 2 Acquisition among HIV-1–Infected Adults Treated with Tenofovir Disoproxyl Fumarate as Part of Combination Antiretroviral Therapy: Results from the ACTG A5175 PEARLS Study. J. Infect. Dis. 2017, 215, 907–910.

37. DeJong C, Spinelli MA, Okochi H, Gandhi M. Tenofovir-based PrEP for COVID-19: an untapped opportunity? AIDS. 2021;35(9):1509–1511.

38. Elfiky, A.A. Ribavirin, Remdesivir, Sofosbuvir, Galidesivir, and Tenofovir against SARS-CoV-2 RNA dependent RNA polymerase (RdRp): A molecular docking study. Life Sci. 2020, 253, 117592.

39. >Edited Udofia, I.A.; Gbayo, K.O.; Oloba-Whenu, O.A.; Ogunbayo, T.B.; Isanbor, C. In silico studies of selected multi-drug targeting against 3CLpro and nsp12 RNA-dependent RNA-polymerase proteins of SARS-CoV-2 and SARS-CoV. Netw. Model. Anal. Health Inform. Bioinform. 2021, 10, 1–12.

40. Poustforoosh, A.; Hashemipour, H.; Tüzün, B.; Pardakhty, A.; Mehrabani, M.; Nematollahi, M.H. Evaluation of potential anti-RNA-dependent RNA polymerase (RdRP) drugs against the newly emerged model of COVID-19 RdRP using computational methods. Biophys. Chem. 2021, 272, 106564.

41. Tiwari, V. Denovo designing, retro-combinatorial synthesis, and molecular dynamics analysis identify novel antiviral VTRM1.1 against RNA-dependent RNA polymerase of SARS CoV2 virus. Int. J. Biol. Macromol. 2021, 171, 358–365.

42. Edited Grahl, M.V.; Alcará, A.M.; de Souza, O.N.; Ligabue-Braun, R.; Perin, A.P.A.; Moro, C.F.; Pinto, É.S.; Feltes, B.C.; Ghilardi, I.M.; Rodrigues, F.V.; et al. Evaluation of drug repositioning by molecular docking of pharmaceutical resources available in the Brazilian healthcare system against SARS-CoV-2. Inform. Med. Unlocked 2021, 23, 100539.

43. Copertino, D.C., Jr.; Lima, B.C.C.; Duarte, R.R.R.; Powell, T.R.; Ormsby, C.E.; Wilkin, T.; Gulick, R.M.; Rougvie, M.D.M.; Nixon, D.F. Antiretroviral drug activity and potential for pre-exposure prophylaxis against COVID-19 and HIV infection. J. Biomol. Struct. Dyn. 2021, 2021, 1–14.

44. Hasan, K.; Kamruzzaman, M.; Bin Manjur, O.H.; Mahmud, A.; Hussain, N.; Alam Mondal, M.S.; Hosen, I.; Bello, M.; Rahman, A. Structural analogues of existing anti-viral drugs inhibit SARS-CoV-2 RNA dependent RNA polymerase: A computational hierarchical investigation. Heliyon 2021, 7, e06435.

45. Dallocchio, R.N.; Dessì, A.; De Vito, A.; Delogu, G.; Serra, P.A.; Madeddu, G. Early combination treatment with existing HIV antivirals: An effective treatment for COVID-19? Eur. Rev. Med. Pharmacol. Sci. 2021, 25, 2435–2448.

46. Salpini, R.; Alkhatib, M.; Costa, G.; Piermatteo, L.; Ambrosio, F.A.; Di Maio, V.C.; Scutari, R.; Duca, L.; Berno, G.; Fabeni, L.; et al. Key genetic elements, single and in clusters, underlying geographically dependent SARS-CoV-2 genetic adaptation and their impact on binding affinity for drugs and immune control. J. Antimicrob. Chemother. 2021, 76, 396–412.

47. Shah, B.; Modi, P.; Sagar, S.R. In silico studies on therapeutic agents for COVID-19: Drug repurposing approach. Life Sci. 2020, 252, 117652.

48. Edited Rahman, M.R.; Banik, A.; Chowdhury, I.M.; Sajib, E.H.; Sarkar, S. Identification of potential antivirals against SARS-CoV-2 using virtual screening method. Inform. Med. Unlocked 2021, 23, 100531.

49. Feng, Z.; Chen, M.; Xue, Y.; Liang, T.; Chen, H.; Zhou, Y.; Nolin, T.D.; Smith, R.B.; Xie, X.-Q. MCCS: A novel recognition pattern-based method for fast track discovery of anti-SARS-CoV-2 drugs. Brief. Bioinform. 2021, 22, 946–962.

50. Yun, Y.; Song, H.; Ji, Y.; Huo, D.; Han, F.; Li, F.; Jiang, N. Identification of therapeutic drugs against COVID-19 through computational investigation on drug repurposing and structural modification. J. Biomed. Res. 2020, 34, 458–469.

51. Toor, H.G.; Banerjee, D.I.; Rath, S.L.; Darji, S.A. Computational drug re-purposing targeting the spike glycoprotein of SARS-CoV-2 as an effective strategy to neutralize COVID-19. Eur. J. Pharmacol. 2021, 890, 173720.

52. Del Amo, J.; Polo, R.; Moreno, S.; Díaz, A.; Martínez, E.; Arribas, J.R.; Jarrín, I.; Hernán, M.A. Incidence and Severity of COVID-19 in HIV-Positive Persons Receiving Antiretroviral Therapy. Ann. Intern. Med. 2020, 173, 536–541.

53. del Amo, J.; Polo, R.; Moreno, S.; Díaz, A.; Martínez, E.; Arribas, J.R.; Jarrín, I.; Hernán, M.A. Antiretrovirals and Risk of COVID-19 Diagnosis and Hospitalization in HIV-Positive Persons. Epidemiology 2020, 31, e49–e51.

54. Jean-Jacques Parienti, Thierry Prazuck, Laure Peyro-Saint-Paul, Anna Fournier, Cécile Valentin, Sylvie Brucato, Renaud Verdon, Aymeric Sève, Mathilda Colin, Fabien Lesne, Jérome Guinard, Meriadeg Ar Gouilh, Julia Dina, Astrid Vabret, Laurent Hocqueloux. Effect of Tenofovir Disoproxil Fumarate and Emtricitabine on nasopharyngeal SARS-CoV-2 viral load burden amongst outpatients with COVID-19: A pilot, randomized, open-label phase 2 trial. EClinicalMedicine 2021:100993.

